# Chronic oxytocin administration stimulates the endogenous oxytocin system: an RCT in autistic children

**DOI:** 10.1101/2023.06.06.23291017

**Authors:** Matthijs Moerkerke, Nicky Daniels, Laura Tibermont, Tiffany Tang, Margaux Evenepoel, Stephanie Van der Donck, Edward Debbaut, Jellina Prinsen, Viktoria Chubar, Stephan Claes, Bart Vanaudenaerde, Lynn Willems, Jean Steyaert, Bart Boets, Kaat Alaerts

**Author notes:** **Corresponding author:** Kaat Alaerts. KU Leuven, Neuromodulation Laboratory, Research Group for Neurorehabilitation, Tervuursevest 101 box 1501, 3001 Leuven, Belgium. shared first authors (Matthijs Moerkerke, Nicky Daniels). shared last authors (Bart Boets, Kaat Alaerts).

## Abstract

**Background:** Clinical efficacy of chronic intranasal administration of oxytocin is increasingly explored in autism spectrum disorder (ASD), but to date, little is known regarding its biological effects and in particular how chronic administration regimes impact endogenous oxytocinergic function.

**Methods:** To fill this gap, this double-blind, randomized, placebo-controlled study explored chronic oxytocin administration effects on endogenous salivary oxytocin levels and oxytocin receptor gene (*OXTR*) epigenetics (DNA methylation) in 8-to-12-year-old children with ASD (n = 79, 16 females). Biological sampling was performed at baseline (pre-treatment), immediately (24 hours) after the four-week oxytocin administration period (12 IU, twice daily) and at a follow-up session, four weeks after the last nasal spray administration.

**Results:** Compared to placebo, children receiving the oxytocin nasal spray displayed significantly higher salivary oxytocin levels 24 hours after the last oxytocin nasal spray administration, but no longer at the four-week follow up session. Regarding epigenetics, oxytocin-induced reductions in *OXTR* methylation were observed, reflecting a facilitation of oxytocin receptor expression in the oxytocin, compared to the placebo group. Notably, heightened oxytocin levels post-treatment were significantly associated with reduced *OXTR* DNA methylation and improved feelings of secure attachment.

**Conclusion:** Four weeks of chronic oxytocin administration stimulated the endogenous oxytocinergic system in children with ASD, as evidenced by increased salivary oxytocin levels and reduced *OXTR* DNA methylation (indicating increased receptor expression).

## Introduction

Autism spectrum disorder (ASD) is a neurodevelopmental condition, characterized by repetitive and restrictive behaviours and socio-communicative difficulties for which limited therapeutic options exist to date (American Psychiatric Association, 2013). The past decade, intranasal administration of oxytocin is increasingly explored as a new approach to facilitate social development and reduce disability associated with ASD (Ooi et al., 2017). However, while initial single-dose administration studies yielded promising acute effects of oxytocin on pro-social behaviour (see review (Ooi et al., 2017)), subsequent multiple-dose, chronic administration studies (i.e. administrating the oxytocin nasal spray over a course of multiple weeks) have yielded a more mixed pattern of results, with some studies demonstrating beneficial outcomes, while others did not (Anagnostou et al., 2012; Bernaerts et al., 2020; Daniels et al., 2023; Le et al., 2022; Sikich et al., 2021).

In light of the growing number of studies demonstrating a limited bioavailability of naturally occurring oxytocin in autistic children (Evenepoel et al., 2022; John & Jaeggi, 2021; Moerkerke, Peeters, et al., 2021), it is crucial to gain deeper insights into how exogenous administration can impact the functioning of the endogenous oxytocin system. This is important, considering that biological changes likely underlie oxytocin-induced behavioural-clinical outcomes (Parker et al., 2017) and therefore allow delineating possible biological mechanisms of inter-individual variation in clinical treatment responses.

To date, insights into how exogenous administration impacts endogenous oxytocinergic signalling have predominantly emerged from single-dose administration studies, showing elevated salivary oxytocin levels up to 2 and even 7 hours after administration in autistic and non-autistic populations (Daughters et al., 2015; Huffmeijer et al., 2012; Procyshyn et al., 2020; Quintana et al., 2018; Riem et al., 2019; van IJzendoorn et al., 2012; Weisman et al., 2012). Considering that the half-life of oxytocin is only a few minutes in blood plasma and 20 minutes in cerebrospinal fluid (Jurek & Neumann, 2018), sustained high levels of salivary oxytocin likely reflect an upregulation of endogenous oxytocin release induced by its acute exogenous administration. This notion is supported by a recent chronic administration study, examining the effect of a four-week course of daily oxytocin administrations on salivary oxytocin levels in autistic adult men (Alaerts et al., 2021). Here, elevated salivary oxytocin levels were shown up to four weeks after cessation of the nasal spray administration period, indicating a self-perpetuating elevation of oxytocin levels through a feed-forward triggering of its own release, in line with the notion of a ‘positive spiral of oxytocin release’ as suggested before by De Dreu (2012). More research is needed, however, to understand the impact of chronic oxytocin administration on its endogenous production, especially in autistic children, considering that oxytocin can preferably exert its therapeutic potential within early life developmental windows.

Aside assessments of circulating oxytocin, also variations in (epi)genetic modifications of the oxytocin receptor gene (*OXTR*) are considered important markers of endogenous oxytocinergic function. DNA methylation (DNAm) is one of the most extensively studied epigenetic mechanisms involved in the regulation of gene transcription, with increased DNAm frequency found to be associated to decreased gene transcription, and therefore less receptor expression and availability (Kusui et al., 2001). While initial studies have linked increased *OXTR* DNAm to social difficulties and traits associated with ASD (Moerkerke, Bonte, et al., 2021), insight in whether (chronic) oxytocin administration impacts *OXTR* DNAm and whether these changes relate to altered oxytocin levels remains currently unexplored.

The current study aims to fill this gap, exploring the biological effects of a four-week course of chronic oxytocin administration on oxytocinergic function as assessed using salivary oxytocin levels and *OXTR* DNAm in school-aged children with ASD.

## Materials & methods

### Study design

A double-blind, randomized, placebo-controlled study with parallel design was conducted at the Leuven University Hospital (Belgium) to assess the effects of four weeks of intranasal oxytocin administration (12IU twice daily) on endogenous levels of salivary oxytocin and *OXTR* DNAm in school-aged children with ASD (see **Fig. 1** for the CONSORT flow diagram visualizing the number of biological samples collected and analysed for each assessment session). Saliva oxytocin and DNA samples were collected at baseline, prior to nasal spray administration (T0); immediately after the four-week administration period, at least 24 hours after the last nasal spray administration (T1); and at a follow-up session, four weeks after cessation of the daily administrations (T2).

**Figure 1.**
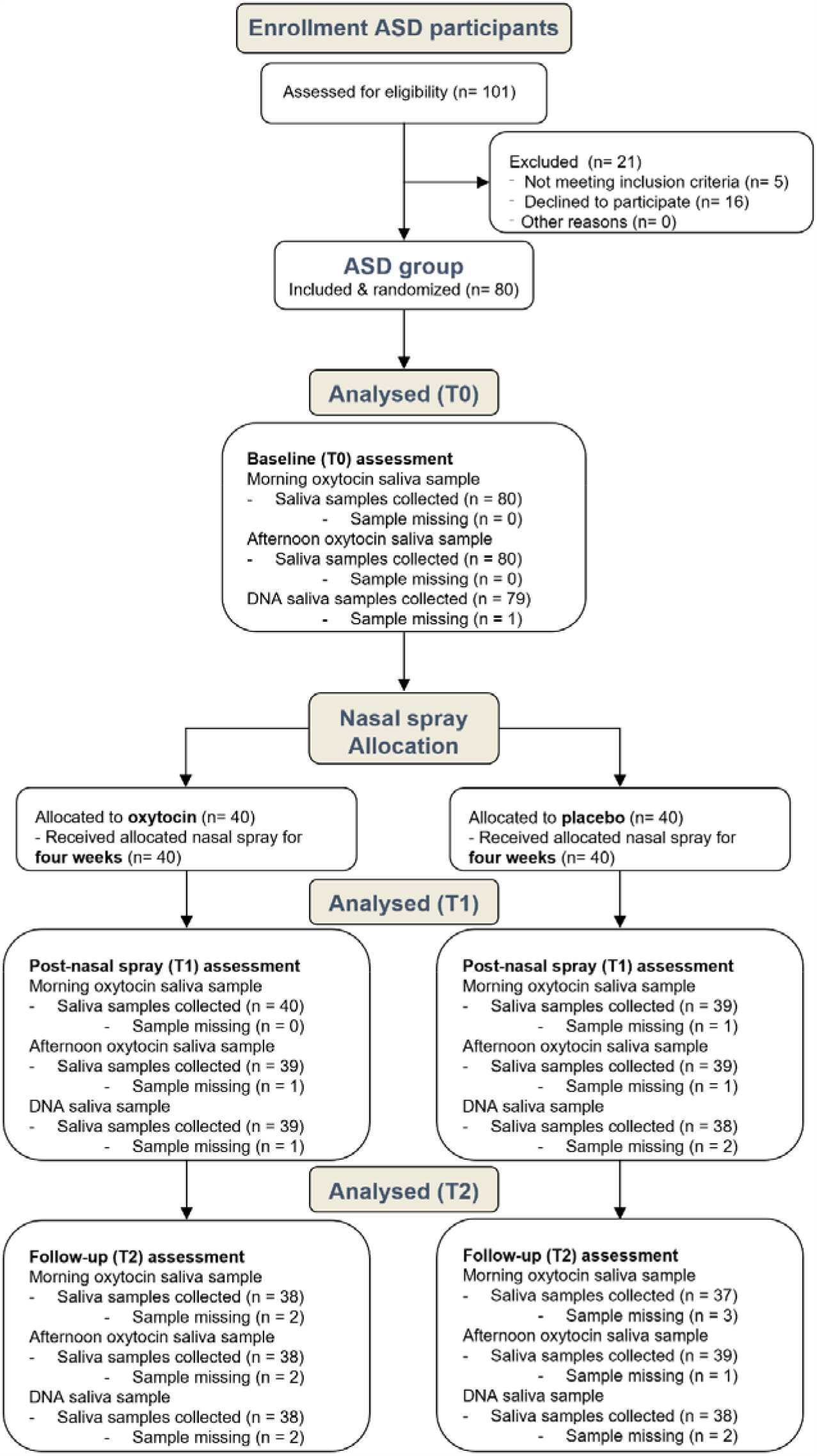
CONSORT flow diagram. Participants were assessed for eligibility prior to the baseline session (T0) and after the session allocated to receive either oxytocin or placebo nasal sprays for a four week period of twice daily nasal spray administration. Saliva oxytocin (morning and afternoon) and DNA samples were collected at baseline (T0), immediately after the four-week administration period (T1; at least 24 hours after the last nasal spray administration), and at a follow-up session, four weeks after cessation of the daily administrations (T2). As outlined, for some participants, saliva samples were missing at one or more assessment sessions due to participant being unable or forgetful to bring the sample or due to discontinuation of the study by the participant.

All study procedures and consent forms were approved by the local Ethics Committee at the KU Leuven (S61358) in accordance with Declaration of Helsinki. Saliva samples were collected in the context of a larger protocol including clinical-behavioural, neural and neurophysiological assessments as registered at the European Clinical Trial Registry (EudraCT 2018-000769-35) and the Belgian Federal Agency for Medicines and Health products (see Supplementary methods).

### Participants

Children with a formal diagnosis of ASD were recruited through the Leuven Autism Expertise Centre between July 2019 and January 2021 (see **Fig. 1** for the CONSORT flow diagram visualizing the number of included participants). Participants randomized to receive oxytocin or placebo nasal sprays did not differ in terms of baseline symptom characteristics (Autism Diagnostic Observation Schedule, ADOS-2, (Lord et al., 2012); Social Responsiveness Scale-Children, SRS-2, (Constantino & Gruber, 2012)), intelligence quotients (IQ; WISC-V-NL, (Wechsler, 2018)), biological sex or age, see **Table 1**. Detailed information regarding inclusion criteria and participant screening is provided in Supplementary methods.

**Table 1.**
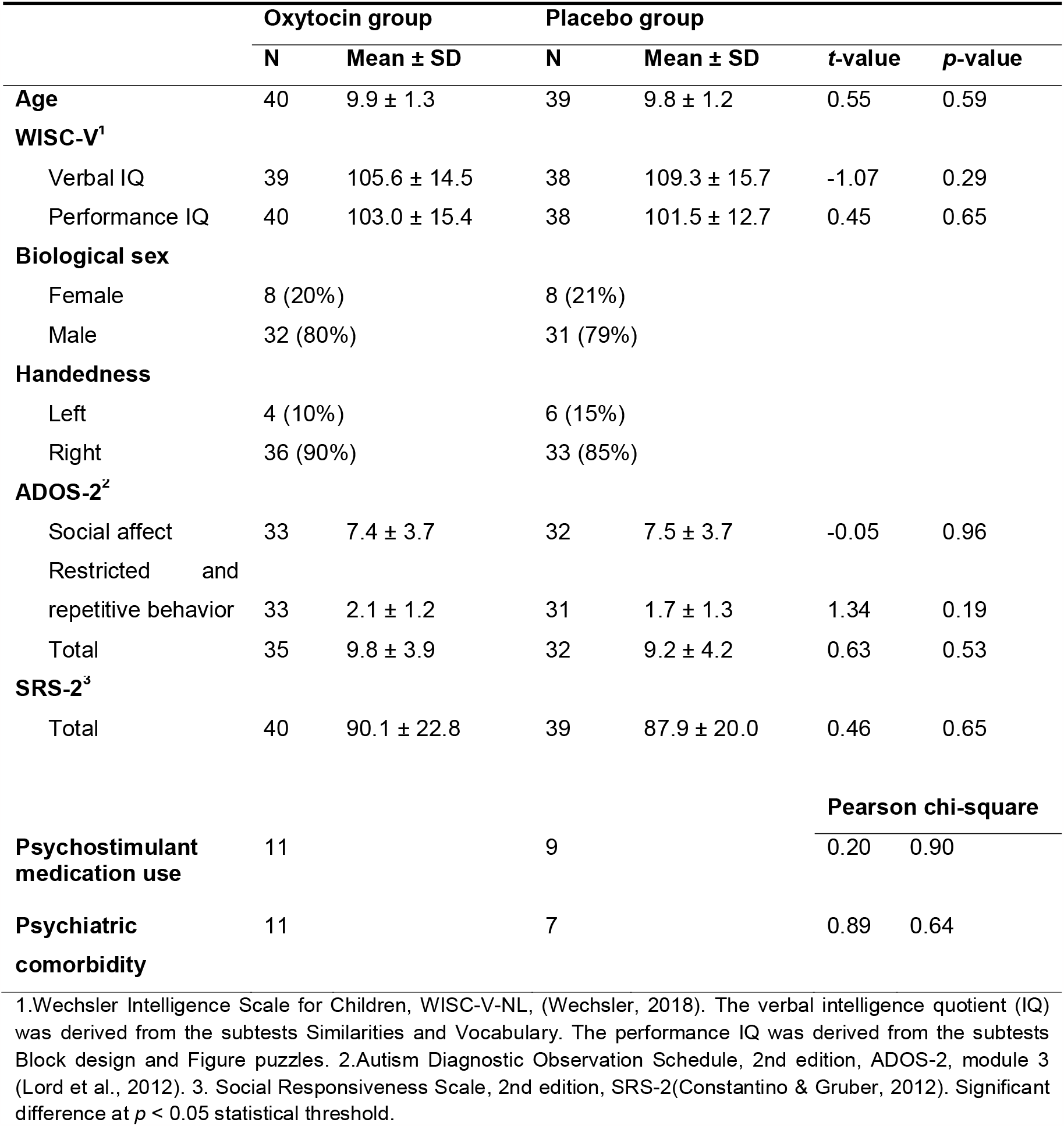
Demographic and clinical characteristics of children randomized to receive oxytocin or placebo. Mean baseline scores, from participants completing at least one post-nasal spray session, are listed separately for each nasal spray administration group. T- and p-values correspond to independent sample t-tests assessing between-group differences in baseline scores.

### Nasal spray administration

Participants received oxytocin (Syntocinon®, Sigma-tau) or placebo nasal sprays for four weeks, administered twice daily with six puffs (three per nostril) of 12 IU in the morning and six puffs of 12 IU in the afternoon (after school), resulting in a daily dose of 24 IU. On day 28, i.e. the last day before the post-nasal spray administration assessment (T1), participants withheld the afternoon spray, to allow a window of 24 hours between the last nasal spray and the salivary sampling. All research staff conducting the trial, participants and their parents were blinded to nasal spray allocation. More information regarding nasal spray randomization, compliance and adverse event screening is provided in Supplementary methods.

### Assessment of salivary oxytocin levels

Oxytocin levels were assessed via saliva samples acquired at each assessment session (T0, T1, T2), at two time points: (i) a morning sample, acquired at home, within 30 min after awakening and before breakfast; and (ii) an afternoon sample, acquired at the Leuven University hospital. Salivary samples were collected using Salivette cotton swabs (Sarstedt AG & Co., Germany) and analysed using a commercial enzyme immunoassay oxytocin ELISA kit (Enzo Life Sciences, Inc., USA) in accordance with the manufacturer’s instructions. Sample concentrations (100 μl/well) were calculated conform plate-specific standard curves.

### Assessment of *OXTR* DNA methylation levels

To assess variations in DNAm of *OXTR* (hg19, chr3:8,810,729-8,810,845), salivary samples were obtained via the Oragene DNA sample collection kit (DNA Genotek Inc., Canada) at the Leuven University hospital at T0, T1, and T2. DNAm was assessed at three CpG sites of *OXTR* that have been shown to be impacted in ASD (i.e., -934, -924 and -914; see (Moerkerke, Bonte, et al., 2021)). More detailed information regarding the DNA collection procedures and analyses are provided in Supplementary methods.

### Data handling and statistical procedures

Detailed information on saliva sample availability and data handling is outlined in Supplementary methods. To assess oxytocin-induced effects, salivary oxytocin levels were subjected to mixed-effects analyses of variances and Bonferroni-corrected post-hoc tests, with the factors ‘*nasal spray*’ (oxytocin, placebo) and ‘*assessment session*’ (T1, T2) as fixed effects, and the factor ‘*subject*’ as random effect. To correct for variance in the individuals’ baseline T0 scores, baseline values prior to nasal administration were included as a covariate in the model. Separate models were constructed for the morning and afternoon samples.

Oxytocin-induced effects in *OXTR* DNAm were analysed using similar mixed models, i.e., with the fixed factors ‘*nasal spray*’ (oxytocin, placebo) and ‘*assessment session*’ (T1, T2), the random factor ‘*subject*’ and baseline (T0) scores modelled as covariate. Separate models were constructed for each CpG site (−934, -924 and -914).Next, Spearman correlation analyses were performed to examine possible associations between salivary oxytocin levels (averaged across morning and afternoon) and *OXTR* DNAm assessed within the oxytocin nasal spray administration group at T1 and T2. Since prior work has evidenced a link between variations in oxytocin levels and behaviour (e.g., Alaerts et al., 2019), associations between salivary oxytocin levels and obtained clinical-behavioural assessments were also performed (see **table S2** for a detailed outlining of the obtained questionnaires). In short, as outlined in Daniels et al., (2023), children receiving oxytocin did not – as a group – display stronger improvements in parent-reported social functioning (assessed using the SRS-2), compared to children receiving the placebo nasal spray. Also for secondary outcomes assessing parent-reported repetitive behaviours (Repetitive Behavior Scale-Revised, RBS-R, (Bodfish et al., 2000)) and anxiety scores (Screen for Child Anxiety Related Emotional Disorders, SCARED-NL, (Muris et al., 2007)), and self-reported attachment (Attachment Style Classification Questionnaire, ASCQ, (Finzi et al., 2000)), no oxytocin-specific improvements were observed. To understand possible inter-individual variations in clinical treatment responses on these outcomes, exploratory examinations were performed to discern whether individual variation in oxytocin-induced effects in endogenous oxytocinergic function may relate to variations in clinical effects. All statistical analyses were executed with Statistica version 14 (Tibco Software Inc.).

## Results

### Effect of chronic oxytocin administration on salivary oxytocin levels

#### Morning oxytocin levels

The main effects of *nasal spray* (F(1,74) = 14.17; p < .001; *η*_*p*_^2^ = .17) and *assessment session* were significant (F(1,74) = 13.29; p < .001; *η*_*p*_^2^ = .16). Yet, these need to be interpreted in the context of the significant *nasal spray x assessment session* interaction effect, indicating that morning oxytocin levels were significantly augmented in the oxytocin group, compared to the placebo group (F(1,74) = 15.71; p < .001; *η*_*p*_^2^), but only at the T1 post assessment (p_Bonferroni_ < .001), not at the T2 four-week follow-up (p_Bonferroni_ > .05), see **Fig. 2A**.

**Figure 2.**
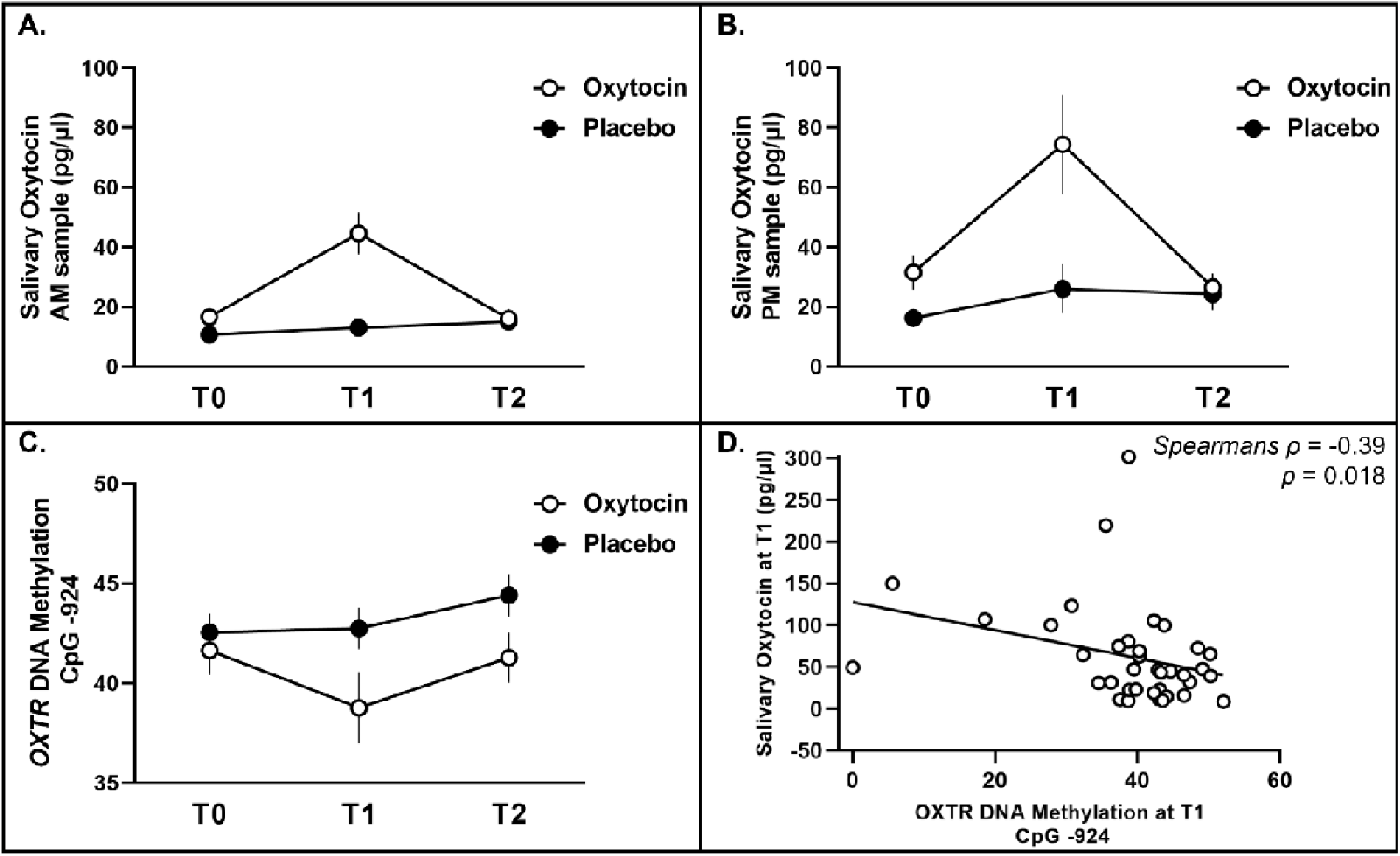
Effect of chronic oxytocin administration on salivary oxytocin levels and *OXTR* DNAm levels. Salivary oxytocin levels are visualised for each nasal spray group (oxytocin and placebo) at each assessment session: baseline (T0), ≥24 hours post-nasal spray (T1) and four weeks post-nasal spray (T2). **Panel A** shows the morning (AM sample) oxytocin levels and **panel B** the afternoon (PM sample) oxytocin levels. **Panel C** visualises the salivary *OXTR* DNAm levels at CpG -924 for each nasal spray group (oxytocin and placebo) at each assessment session (T0, T1 and T2). **Panel D** visualises the association of salivary oxytocin levels (averaged over the morning and afternoon samples) with the salivary *OXTR* DNAm levels in CpG -924 in the oxytocin group at T1. Vertical bars denote standard errors.

#### Afternoon oxytocin levels

An overall similar pattern of results was identified for afternoon oxytocin levels, indicating a significant augmentation in the oxytocin group compared to the placebo group (F(1,71) = 6.69; p = .012; *η*_*p*_^2^ = .08), at the T1 post assessment (*p*_*Bonferroni*_ = .001), not at the T2 follow-up (*p* > .05), see **Fig. 2B**.

### Effect of chronic oxytocin administration on salivary *OXTR* DNAm

#### CpG site -924

A significant main effect of *nasal spray* was identified (F(1,70) = 7.76; *p* = .007; *η*_*p*_^2^ = .10), indicating that DNAm of CpG site -924 was significantly reduced in the oxytocin group, compared to the placebo group, see **Fig. 2C**. The absence of a *spray × assessment session* interaction (F(1,70) = .90; p = .346; *η*_*p*_^2^ = .01) indicates that the main effect of nasal spray was evident across both the T1 post and T2 follow-up assessment. The significant main effect of *assessment session* indicates overall higher DNAm at the T2 assessment compared to the T1 assessment (F(1,70) = 6.76; *p* = .011; *η*_*p*_^2^ = .085).

#### CpG site -914 and -934

No significant main nor interaction effects were identified for CpG site -914 and -934 (all *p* > .05), indicating no differential modulation of DNAm at these sites after oxytocin or placebo nasal spray administration, see **Fig. S1**.

### Associations of oxytocin levels with *OXTR* DNAm

Notably, in the oxytocin group, a significant association was evident between salivary oxytocin levels post-nasal spray, at T1, and DNAm of CpG site -924 (*ρ* = -.39; *p* = .018), indicating that children of the oxytocin group displaying higher oxytocin levels at T1 also showed a stronger reduction in DNAm of CpG site -924 (reflecting higher oxytocin-receptor expression), see **Fig. 2D**. No significant associations were evident at the T2 session (*ρ* = .09; *p* = .577).

### Associations with behavioural-clinical reports

A significant relationship was identified between salivary oxytocin levels assessed from the oxytocin group at T1 and self-reports of secure attachment towards peers (ASCQ), indicating that children of the oxytocin group displaying higher oxytocin levels at T1 also showed higher reports of secure attachment at T1 (*ρ* = .35; *p* = .030), see **Fig. 3A**. Moderate (but non-significant) opposite relationships were evident for self-reported attachment avoidance (*ρ* = -.30; *p* = .070, **Fig. 3B**) and parent-reports of social responsiveness (SRS-2; *ρ* = -.31; *p* = .062, **Fig. 3C**) and anxiety (SCARED-NL; *ρ* = -.32; *p* = .053, **Fig. 3D**) all indicating that improved clinical presentations at T1 were associated with higher T1 oxytocin levels. No significant associations were identified between levels of DNAm of CpG site -924 and the abovementioned clinical scales.

**Figure 3.**
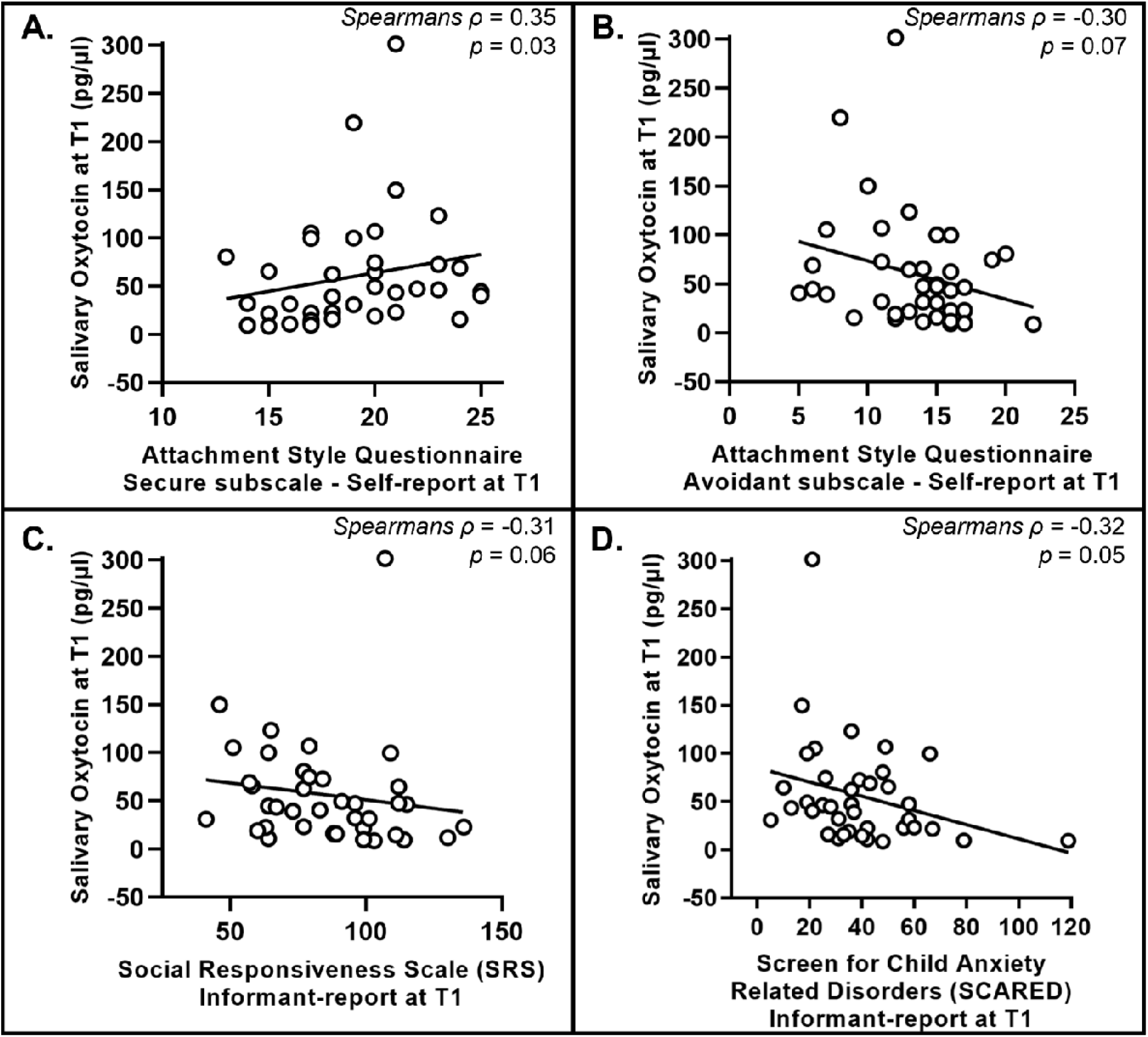
Associations of oxytocin levels with questionnaires assessing attachment style, social responsivity and anxiety. The association of salivary oxytocin levels with questionnaire scores at the post oxytocin nasal sprays session (T1) are visualised. **Panel A** shows the association with secure-attachment (Attachment Style Classification Questionnaire, ASCQ). **Panel B** shows the association with avoidant-attachment (ASCQ). **Panel C** shows the association with social responsiveness (Social Responsiveness Scale, SRS). **Panel D** shows the association with anxiety scores (Screen for Child Anxiety Related Emotional Disorders, SCARED).

## Discussion

This study examined the impact of a four-week chronic oxytocin administration regime on endogenous oxytocinergic function in autistic children. Salivary oxytocin levels were reliably increased 24 hours after the last oxytocin nasal spray, but no longer at the follow-up session, four weeks after cessation of the oxytocin administrations. Reduced *OXTR* DNAm levels were also observed, suggesting a facilitation in oxytocin-receptor expression in the oxytocin, compared to the placebo group, up to four weeks after the last nasal spray administration. Increased oxytocin levels were significantly associated with reduced *OXTR* DNAm levels, as well as with improved clinical presentations on self-reported feelings of secure peer attachment. These findings provide evidence that the chronic exogenous administration induced stimulation of endogenous oxytocinergic system in children with ASD.

Although the exact physiological mechanisms remain unclear, prior research showed that oxytocin-receptor binding promotes secretion of additional oxytocin and affects DNA transcriptional activation/repression (Gulliver et al., 2019; Jurek & Neumann, 2018; Neumann et al., 1996). It is therefore plausible that the chronic exogenous oxytocin administration exerted a facilitatory impact on the release of endogenous oxytocin, as well as stimulated transcription of the oxytocin receptor through epigenetic modifications of the *OXTR* gene as observed in the current cohort. Since epigenetic mechanisms constitute a direct pathway by which environmental changes (e.g. increased oxytocin availability) may impact DNA transcriptional activation/repression, the observed changes in DNAm of *OXTR* likely reflect an important biological pathway through which chronic oxytocin administration regimes may induce long-lasting neurobiological and possibly associated clinical-behavioural effects. In short, the elevated oxytocin availability may have induced a feed-forward mechanism of the endogenous oxytocinergic function by increasing its own release, as well as prompting more sustained epigenetic *OXTR* modifications.

The stimulating effect of chronic oxytocin administration on endogenous oxytocin levels in autistic children, observed at T1 but not at T2, is partly in line with prior observations in adults with ASD (Alaerts et al., 2021). In the latter study, increases in endogenous oxytocin levels persisted up to four weeks after cessation of the chronic nasal spray administrations. Developmental differences of the oxytocinergic system have already been evidenced in prior studies, indicating lower endogenous oxytocin levels in autistic children, but not adults (Moerkerke, Peeters, et al., 2021). Perhaps in autistic children, more so than in adults, homeostatic responses upon cessation of the chronic oxytocin administration occur faster, thereby reducing heightened levels of circulating oxytocin more rapidly to initial baseline levels. Alternatively, adults (as compared to children) might adapt their social interactive behaviour more readily upon receiving oxytocin administrations, i.e., through increasingly engaging in and positively experiencing of socially stimulating contexts. According to the positive spiral of oxytocin release, this increase in (positive) social experiences may have further facilitated oxytocin’s endogenous release, also in the long-term, as observed up to four-weeks after cessation of the nasal spray administration regime in adults (Alaerts et al., 2021), but not children in the current cohort. In light of potential treatment applications, this notion may highlight the importance of combining oxytocin administration with social stimulation to perpetuate the endogenous oxytocin feedforward-loop (De Dreu, 2012), however future research is needed to explore the differential impact of this combination.

It is noteworthy that higher levels of endogenous oxytocin observed post-treatment were linked to better self-reported feelings of secure attachment. Additionally, there was a trend towards a reduced self-reported attachment avoidance, as well as parent-reported social difficulties and anxiety. A large body of literature links variations in circulating oxytocin to variations in behavioural manifestations, including prosociality and attachment-related constructs (Ferreira & Osório, 2022; Marazziti et al., 2006; Valstad et al., 2017). Over the past decades, the role of the oxytocinergic system in inter-personal bonding and formation and maintenance of secure attachment bonds was highlighted. Our study provides a first demonstration of the link between elevated oxytocin levels and a better clinical presentation of secure attachment in autistic children, supporting the notion that also in autistic children heightened oxytocin may facilitate a sense of (social) security and safety (Carter, 2014; Porges, 2011).

In the current study, salivary sampling was adopted for both oxytocin level and *OXTR* DNAm characterizations, as these assessments were most feasible in a paediatric population. While correlations have been demonstrated between salivary and central (cerebrospinal fluid) oxytocin levels (Martin et al., 2018), salivary oxytocin may constitute only a proxy of central oxytocinergic function. Future studies are therefore warranted to examine whether the currently reported effects - both in terms of salivary oxytocin and epigenetic modification – would hold also for other biological material, including blood and cerebrospinal fluid. Further, considering the tight age range of our school-aged children with ASD, and a predominant sampling from boys with ASD, it should be explored whether the observed effects will generalize across different age ranges and in samples with a larger representation of girls.

To conclude, four weeks of chronic oxytocin administration was shown to stimulate the endogenous oxytocinergic system in autistic children, as evidenced by increased salivary oxytocin levels and associated decreased *OXTR* DNAm levels (reflecting increased oxytocin-receptor expression). Furthermore, elevated oxytocin levels post-treatment were associated with a better clinical presentation, indicating an important mechanistic link between oxytocin’s chronic biological and clinical-behavioural effects.

## Data Availability

All data produced in the present study are available upon reasonable request to the authors

## Author Contributions

**M.M**.: conceptualization, methodology, investigation, data curation, validation, writing—original draft, writing—review and editing, visualization, project administration. **N.D**.: conceptualization, investigation, data curation, validation, writing—review and editing, project administration. **L.T**.: methodology, investigation, data curation, validation, writing—review and editing. **T.T**.: methodology, investigation, data curation, validation, writing—review and editing. **M.E**.: methodology, writing—review and editing. **S.VdD**.: data curation, validation, writing—review and editing. **E.D**.: data curation, validation, writing—review and editing. **J.P**.: data curation, validation, writing—review and editing. **V.C**.: methodology, writing—review and editing. **S.C**.: methodology, writing—review and editing. **B.V**.: methodology, writing—review and editing. **L.W**.: methodology, writing—review and editing. **J.S**.: supervision, validation, writing—review and editing, funding acquisition. **B.B**.: supervision, conceptualization, validation, writing—review and editing, funding acquisition. **K.A**.: supervision, conceptualization, methodology, validation, writing—original draft, writing—review and editing, visualization, project administration, funding acquisition. All authors have read and agreed to the published version of the manuscript.

## References

Alaerts, K., Bernaerts, S., Vanaudenaerde, B., Daniels, N., & Wenderoth, N. (2019). Amygdala– Hippocampal Connectivity Is Associated With Endogenous Levels of Oxytocin and Can Be Altered by Exogenously Administered Oxytocin in Adults With Autism. Biological Psychiatry: Cognitive Neuroscience and Neuroimaging, 4(7), 655–663. https://doi.org/10.1016/j.bpsc.2019.01.008

Alaerts, K., Steyaert, J., Vanaudenaerde, B., Wenderoth, N., & Bernaerts, S. (2021). Changes in endogenous oxytocin levels after intranasal oxytocin treatment in adult men with autism: An exploratory study with long-term follow-up. European Neuropsychopharmacology, 43, 147–152. https://doi.org/10.1016/J.EURONEURO.2020.11.014

American Psychiatric Association. (2013). Diagnostic and statistical manual of mental disorders (5th ed.). In Washington, DC.

Anagnostou, E., Soorya, L., Chaplin, W., Bartz, J., Halpern, D., Wasserman, S., Wang, A. T., Pepa, L., Tanel, N., Kushki, A., & Hollander, E. (2012). Intranasal oxytocin versus placebo in the treatment of adults with autism spectrum disorders: A randomized controlled trial. Molecular Autism, 3(1), 1–9. https://doi.org/10.1186/2040-2392-3-16/TABLES/3

Bernaerts, S., Boets, B., Bosmans, G., Steyaert, J., & Alaerts, K. (2020). Behavioral effects of multiple-dose oxytocin treatment in autism: A randomized, placebo-controlled trial with long-term follow-up. Molecular Autism, 11(1), 1–14. https://doi.org/10.1186/s13229-020-0313-1

Bodfish, J. W., Symons, F. J., Parker, D. E., & Lewis, M. H. (2000). Varieties of repetitive behaviour in autism: comparison to mental retardation. Journal of Autism and Developmental Disorders, 30(3), 237–243. https://doi.org/10.1023/A:1005596502855

Carter, C. S. (2014). Oxytocin pathways and the evolution of human behavior. Annual Review of Psychology, 65, 17–39. https://doi.org/10.1146/annurev-psych-010213-115110

Constantino, J., & Gruber, C. (2012). Social responsiveness scale 2nd. ed: SRS-2. Manual. Western Psychological Services. http://kkjp.devswitch.nl/wp-content/uploads/2018/04/Social-Responsiveness-Scale-SRS.pdf

Daniels, N., Moerkerke, M., Steyaert, J., Bamps, A., Debbaut, E., Prinsen, J., Tang, T., Van Der Donck, S., Boets, B., & Alaerts, K. (2023). Effects of multiple-dose intranasal oxytocin administration on social responsiveness in children with autism: a randomized, placebo-controlled trial. Molecular Autism, 14, 16. https://doi.org/10.1186/s13229-023-00546-5

Daughters, K., Manstead, A. S. R., Hubble, K., Rees, A., Thapar, A., & Van Goozen, S. H. M. (2015). Salivary Oxytocin Concentrations in Males following Intranasal Administration of Oxytocin: A Double-Blind, Cross-Over Study. https://doi.org/10.1371/journal.pone.0145104

De Dreu, C. K. W. (2012). Oxytocin modulates cooperation within and competition between groupslll: An integrative review and research agenda. Hormones and Behavior, 61(3), 419–428. https://doi.org/10.1016/j.yhbeh.2011.12.009

Evenepoel, M., Moerkerke, M., Daniels, N., Chubar, V., Claes, S., Turner, J., Vanaudenaerde, B., Willems, L., Verhaeghe, J., Prinsen, J., Steyaert, J., Boets, B., & Alaerts, K. (2022). Endogenous oxytocin levels in children with autism: Associations with cortisol levels and oxytocin receptor gene methylation. MedRxiv, 2022.12.15.22283521. https://doi.org/10.1101/2022.12.15.22283521

Ferreira, A. C., & Osório, F. de L. (2022). Peripheral oxytocin concentrations in psychiatric disorders – A systematic review and methanalysis: Further evidence. Progress in Neuro-Psychopharmacology and Biological Psychiatry, 117, 110561. https://doi.org/10.1016/J.PNPBP.2022.110561

Finzi, R., Cohen, O., Sapir, Y., & Weizman, A. (2000). Attachment Styles in Maltreated Children: A Comparative Study. Child Psychiatry and Human Development 2000 31:2, 31(2), 113–128. https://doi.org/10.1023/A:1001944509409

Gulliver, D., Werry, E., Reekie, T. A., Katte, T. A., Jorgensen, W., & Kassiou, M. (2019). Targeting the Oxytocin System: New Pharmacotherapeutic Approaches. Trends in Pharmacological Sciences, 40(1), 22–37. https://doi.org/10.1016/J.TIPS.2018.11.001

Huffmeijer, R., Alink, L. R. A., Tops, M., Grewen, K. M., Light, K. C., Bakermans-Kranenburg, M. J., & Van Ijzendoorn, M. H. (2012). Salivary levels of oxytocin remain elevated for more than two hours after intranasal oxytocin administration. Neuroendocrinology Letters, 33(1), 22467107–330112. https://www.nel.edu

John, S., & Jaeggi, A. V. (2021). Oxytocin levels tend to be lower in autistic children: A meta-analysis of 31 studies: Https://Doi.Org/10.1177/13623613211034375, 136236132110343. https://doi.org/10.1177/13623613211034375

Jurek, B., & Neumann, I. D. (2018). The oxytocin receptor: From intracellular signaling to behavior. Physiological Reviews, 98(3), 1805–1908. https://doi.org/10.1152/PHYSREV.00031.2017/ASSET/IMAGES/LARGE/Z9J0021828400013.JPEG

Kusui, C., Kimura, T., Ogita, K., Nakamura, H., Matsumura, Y., Koyama, M., Azuma, C., & Murata, Y. (2001). DNA methylation of the human oxytocin receptor gene promoter regulates tissue-specific gene suppression. Biochemical and Biophysical Research Communications, 289(3), 681– 686. https://doi.org/10.1006/bbrc.2001.6024

Le, J., Zhang, L., Zhao, W., Zhu, S., Lan, C., Kou, J., Zhang, Q., Zhang, Y., Li, Q., Chen, Z., Fu, M., Montag, C., Zhang, R., Yang, W., Becker, B., & Kendrick, K. M. (2022). Infrequent Intranasal Oxytocin Followed by Positive Social Interaction Improves Symptoms in Autistic Children: A Pilot Randomized Clinical Trial. Psychotherapy and Psychosomatics, 1–13. https://doi.org/10.1159/000524543

Lord, C., Rutter, M., Dilavore, P. C., Risi, S., Gotham, K., Bishop, S. L., Luyster, R. J., & Guthrie, W. (2012). ADOS-Autisme diagnostisch observatieschema Handleiding.

Marazziti, D., Dell’Osso, B., Baroni, S., Mungai, F., Catena, M., Rucci, P., Albanese, F., Giannaccini, G., Betti, L., Fabbrini, L., Italiani, P., Del Debbio, A., Lucacchini, A., & Dell’Osso, L. (2006). A relationship between oxytocin and anxiety of romantic attachment. Clinical Practice and Epidemiology in Mental Health, 2(1), 1–6. https://doi.org/10.1186/1745-0179-2-28/FIGURES/1

Martin, J., Kagerbauer, S. M., Gempt, J., Podtschaske, A., Hapfelmeier, A., & Schneider, G. (2018). Oxytocin levels in saliva correlate better than plasma levels with concentrations in the cerebrospinal fluid of patients in neurocritical care. Journal of Neuroendocrinology, 30(5), e12596. https://doi.org/10.1111/JNE.12596

Moerkerke, M., Bonte, M. L., Daniels, N., Chubar, V., Alaerts, K., Steyaert, J., & Boets, B. (2021). Oxytocin receptor gene (OXTR) DNA methylation is associated with autism and related social traits-A systematic review. Research in Autism Spectrum Disorders, 85(April), 101785. https://doi.org/10.1016/j.rasd.2021.101785

Moerkerke, M., Peeters, M., de Vries, L., Daniels, N., Steyaert, J., Alaerts, K., & Boets, B. (2021). Endogenous Oxytocin Levels in Autism—A Meta-Analysis. Brain Sciences 2021, Vol. 11, Page 1545, 11(11), 1545. https://doi.org/10.3390/BRAINSCI11111545

Muris, P., Bodden, D., Hale, W., Birmaher, B., & Mayer, B. (2007). SCARED-NL. Vragenlijst over angst en bang-zijn bij kinderen en adolescenten. Handleiding bij de gereviseerde Nederlandse versie van de Screen for Child Anxiety Related Emotional Disorders. Boom Uitgevers.

Neumann, I., Douglas, A. J., Pittman, Q. J., Russell, J. A., & Landgraf, R. (1996). Oxytocin Released within the Supraoptic Nucleus of the Rat Brain by Positive Feedback Action is Involved in Parturition-Related Events. Journal of Neuroendocrinology, 8(3), 227–233. https://doi.org/10.1046/J.1365-2826.1996.04557.X

Ooi, Y. P., Weng, S. J., Kossowsky, J., Gerger, H., & Sung, M. (2017). Oxytocin and Autism Spectrum Disorders: A Systematic Review and Meta-Analysis of Randomized Controlled Trials. Pharmacopsychiatry, 50(1), 5–13. https://doi.org/10.1055/S-0042-109400/ID/R2016-02-0549-0027

Parker, K. J., Oztan, O., Libove, R. A., Sumiyoshi, R. D., Jackson, L. P., Karhson, D. S., Summers, J. E., Hinman, K. E., Motonaga, K. S., Phillips, J. M., Carson, D. S., Garner, J. P., & Hardan, A. Y. (2017). Intranasal oxytocin treatment for social deficits and biomarkers of response in children with autism. Proceedings of the National Academy of Sciences of the United States of America, 114(30), 8119–8124. https://doi.org/10.1073/pnas.1705521114

Porges, S. W. (2011). The Polyvagal Theory: Neurophysiological Foundations of Emotions, Attachment, Communication, Self-Regulation (Norton Series on Interpersonal Neurobiology). WW Norton & Company.

Procyshyn, T. L., Lombardo, M. V., Lai, M. C., Auyeung, B., Crockford, S. K., Deakin, J., Soubramanian, S., Sule, A., Baron-Cohen, S., & Bethlehem, R. A. I. (2020). Effects of oxytocin administration on salivary sex hormone levels in autistic and neurotypical women. Molecular Autism, 11(1), 1–11. https://doi.org/10.1186/S13229-020-00326-5/FIGURES/3

Quintana, D. S., Westlye, L. T., Smerud, K. T., Mahmoud, R. A., Andreassen, O. A., & Djupesland, P. G. (2018). Saliva oxytocin measures do not reflect peripheral plasma concentrations after intranasal oxytocin administration in men. Hormones and Behavior, 102, 85–92. https://doi.org/10.1016/J.YHBEH.2018.05.004

Riem, M. M. E., van IJzendoorn, M. H., & Bakermans-Kranenburg, M. J. (2019). Hippocampal volume modulates salivary oxytocin level increases after intranasal oxytocin administration. Psychoneuroendocrinology, 101, 182–185. https://doi.org/10.1016/J.PSYNEUEN.2018.11.015

Sikich, L., Kolevzon, A., King, B. H., McDougle, C. J., Sanders, K. B., Kim, S.-J., Spanos, M., Chandrasekhar, T., Trelles, M. D. P., Rockhill, C. M., Palumbo, M. L., Cundiff, A. W., Montgomery, A., Siper, P., Minjarez, M., Nowinski, L. A., Marler, S., Shuffrey, L. C., Alderman, C., … Veenstra-VanderWeele, J. (2021). Intranasal Oxytocin in Children and Adolescents with Autism Spectrum Disorder. The New Engl and Journal of Medicine, 385(16), 1462–1473. https://www.nejm.org/doi/10.1056/NEJMoa2103583

Valstad, M., Alvares, G. A., Egknud, M., Matziorinis, A. M., Andreassen, O. A., Westlye, L. T., & Quintana, D. S. (2017). The correlation between central and peripheral oxytocin concentrations: A systematic review and meta-analysis. Neuroscience and Biobehavioral Reviews, 78(April), 117– 124. https://doi.org/10.1016/j.neubiorev.2017.04.017

van IJzendoorn, M. H., Bhandari, R., van der Veen, R., Grewen, K. M., & Bakermans-Kranenburg, M. J. (2012). Elevated Salivary Levels of Oxytocin Persist More than 7 h after Intranasal Administration. Frontiers in Neuroscience, 6, 174. https://doi.org/10.3389/FNINS.2012.00174

Wechsler, D. (2018). WISC-V-NL. Wechsler Intelligence Scale for Children, Fifth Edition, Dutch version. Amsterdam: Pearson Benelux B.V.

Weisman, O., Zagoory-Sharon, O., & Feldman, R. (2012). Intranasal oxytocin administration is reflected in human saliva. Psychoneuroendocrinology, 37(9), 1582–1586. https://doi.org/10.1016/j.psyneuen.2012.02.014

