## Supplemental Material for "Chronic oxytocin administration stimulates the endogenous oxytocin system: an RCT in autistic children"

* shared first authors

^†^ shared last authors

### Supplementary methods

#### Study design

The collection of saliva samples throughout the course of the clinical trial was part of a larger protocol evaluating the effects of oxytocin administration on clinical-behavioural measures (Daniels et al., 2023), neural measures (Moerkerke et al., 2023) and neurophysiological measures (Alaerts et al., 2023), as registered at the European Clinical Trial Registry (EudraCT 2018-000769-35). Additionally, in Evenepoel et al., (2022), a report on endogenous oxytocin level differences between children with and without ASD, prior to any intervention was disseminated.

#### Participants

Main inclusion criteria comprised a clinical diagnosis of ASD, age (8-12 years old), IQ above 70, no prior oxytocin administrations and native Dutch speaker. The ASD diagnosis was established by a multidisciplinary team (child psychiatrist, psychologist, speech/language pathologist, physiotherapist) based on the criteria of the Diagnostic and Statistical Manual of Mental Disorders, 5th edition (American Psychiatric Association, 2013). Main exclusion criteria comprised a history of any neurological disorder (stroke, concussion, epilepsy etc.), any physical disorder (liver, renal, cardiac pathology) or significant hearing or vision impairments. Only premenarchal girls were included. Seventeen participants had a co-occurring diagnosis of attention deficit/hyperactivity disorder and one of obsessive compulsive disorder. Twenty participants used stimulant medication (e.g. methylphenidate), 13 anti-psychotics (e.g. risperidone), 4 anti-depressants (e.g. sertraline) and 35 used other medication (e.g. sleeping aids, gastro-intestinal medication or nutritional supplements).

#### Nasal spray administration

Using permuted-block randomization, participants were randomly assigned to receive oxytocin (Syntocinon®, Sigma-Tau) or placebo nasal sprays, administered in identical blinded amber 10 ml glass bottles with a white nasal pump (0.05 ml or 2 IU /puff). Preparation, packaging and blinding of the nasal sprays was performed by Heidelberg University Hospital, Germany. Placebo sprays contained all the ingredients used in the active solution except the oxytocin compound. Participants were randomly assigned in a 1:1 ratio, with age, IQ and biological sex balanced across treatment arms (see Table 1). Children and their parents received clear instructions on how to administer the nasal spray (Guastella et al., 2013) through a demonstration together with the experimenter. Two daily doses of 12 IU oxytocin nasal spray or placebo equivalent (3 puffs of 2 IU in each nostril) were administered: 12 IU in the morning and 12 IU in the afternoon, during 28 consecutive days. The last day before the post-nasal spray administration assessment session (T1), participants withheld the afternoon spray, to allow a window of 24 hours between the last nasal spray and the salivary sampling. Compliance was monitored by weighing the disposed and returned nasal sprays, no differences were reported in the amount of administered fluid between the oxytocin and placebo group (see Daniels et al. (2023) for a full report). Potential adverse events were tracked using weekly parent reports and daily parent and child diaries. These revealed no evidence of nasal spray-specific adverse events, see Daniels et al. (2023) for a full report.

#### Assessment of *OXTR* DNA methylation levels

Salivary samples were collected at the end of each study visit using the Oragene DNA sample collection kit (DNA Genotek Inc., Canada), to explore epigenetic variations of the oxytocin receptor gene (*OXTR*; hg19, chr3:8,810,729-8,810,845). Next, extracted DNA (200 ng) was bisulfite-converted following the manufacturer's protocol (EZ-96 DNAm Kit, Zymo Research, Irvine, USA) and stored at −80 °C. DNA methylation (DNAm) levels were determined at three CpG sites (-934, -924 and -914) using Pyrosequencing (Qiagen, Germany) and analysed using Pyromark Q96 software, in accordance with manufacturer’s protocols (Luis Royo et al., 2007). Protocols for the PCR amplification and Pyrosequencing analysis were adopted from (Krol et al., 2019). To amplify the DNA fragment of interest, within the *OXTR*, including the CpG sites -934, -924 and -914, the following PCR primers were used: [*OXTR* Forward: TTG AGT TTT GGA TTT AGA TAA TTA AGG ATT; *OXTR* Reverse: /5Biosg/AC TTA ACA TCA CAT TAA ATA CAA CC]. The following PCR amplification steps were used: Step 1: (95°C/15 min)/1 cycle, Step 2: (94°C/30 s, 58°C/30 s, 72°C/30 s)/50 cycles, Step 3: (72°C/10 min)/1 cycle, Step 4: 4°C hold]. The following sequencing primer was used to read the DNA sequence (*OXTR* Sequencing: AGA AGT TAT TTT ATA ATT TT).

Data handling and statistical procedures

**Salivary oxytocin samples.** From oxytocin saliva samples containing less than 100 µl of material, 50 µl was obtained and diluted to 100 µl, sample concentrations were then multiplied by 2 (T0: 4 AM samples, 1 PM sample; T1: 3 AM samples; T2: 3 AM samples). Further, for participants displaying concentrations below the detection limit, concentrations were set to the lowest detected value across samples (T0: 14 AM samples, 12 PM samples; T1: 14 AM samples, 9 PM samples; T2: 18 AM samples, 9 PM samples). Finally, extreme outliers in oxytocin level data were identified (five inter-quartile ranges (Q3-Q1) below or above the first (Q1), respectively third (Q3) quartile) and recoded to the sample mean (across groups) (T0: 1 AM sample, 1 PM sample; T1: 3 AM samples; T2: 1 AM sample, 1 PM sample). Note that the pattern of results remained qualitatively similar also when removing these outliers.

**Salivary DNAm samples.** Samples where *OXTR* DNAm could not be determined due to insufficient sample quality, were removed from analysis (T0: 2 samples, T1: 2 samples). For the DNAm level data, only one extreme outlier was identified and recoded to the sample mean (across groups), namely for CpG site -934, at T0.

#### Behavioural measures

**Table S1. Detailed description of the adopted behavioural measures.**

| **Outcome measures** | **Construct** | **Type of report** | **Meaning of higher scores** | **Reference** |
| --- | --- | --- | --- | --- |
| **Social Responsiveness Scale-Children**  **(SRS-2)** | Symptom severity | Parent-reported questionnaire | Greater deficits in social responsiveness | Constantino & Gruber, 2012; Roeyers et al., 2015 |
| **Autism Diagnostic Observation Schedule (ADOS-2)** | Symptom severity | Observation | More severe symptoms of autism spectrum disorder | Lord et al., 2012 |
| **Wechsler Intelligence Scale for Children (WISC-V-NL)** | Verbal Intelligence Quotient | Observation | Higher verbal abilities | Wechsler, 2018 |
|  | Performance Intelligence Quotient |  | Higher visual spatial abilities |  |
| **Screen for Child Anxiety Related Emotional Disorders (SCARED-NL)** | Anxiety | Parent-reported questionnaire | Higher risk for anxiety disorders | Muris et al., 2007 |
| **Attachment Style Classification Questionnaire (ASCQ)**  **Anxious**  **Avoidant**  **Secure** | Attachment | Self-reported questionnaire | More anxious, avoidant or secure attachment toward their peers | Finzi et al., 2000 |

### Supplementary results

**Figure S1. Effect of chronic oxytocin administration on *OXTR* DNAm levels at CpG sites -914 and -934.** Visualisation of the salivary *OXTR* DNAm levels at CpG -914 and -934 for each nasal spray group (oxytocin and placebo) at each assessment session (T0, T1 and T2). Vertical bars denote standard errors.

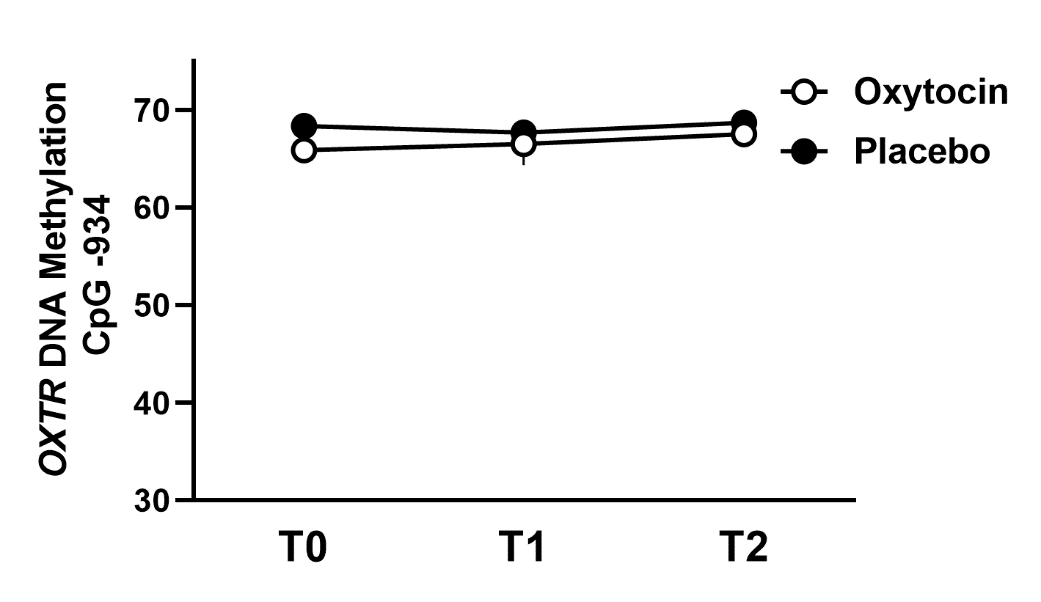

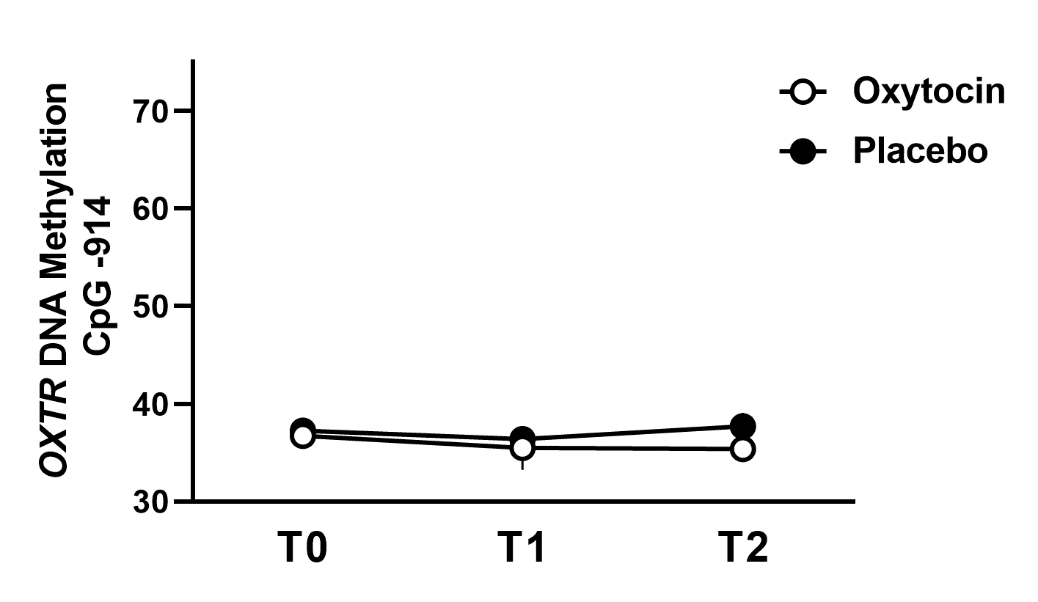

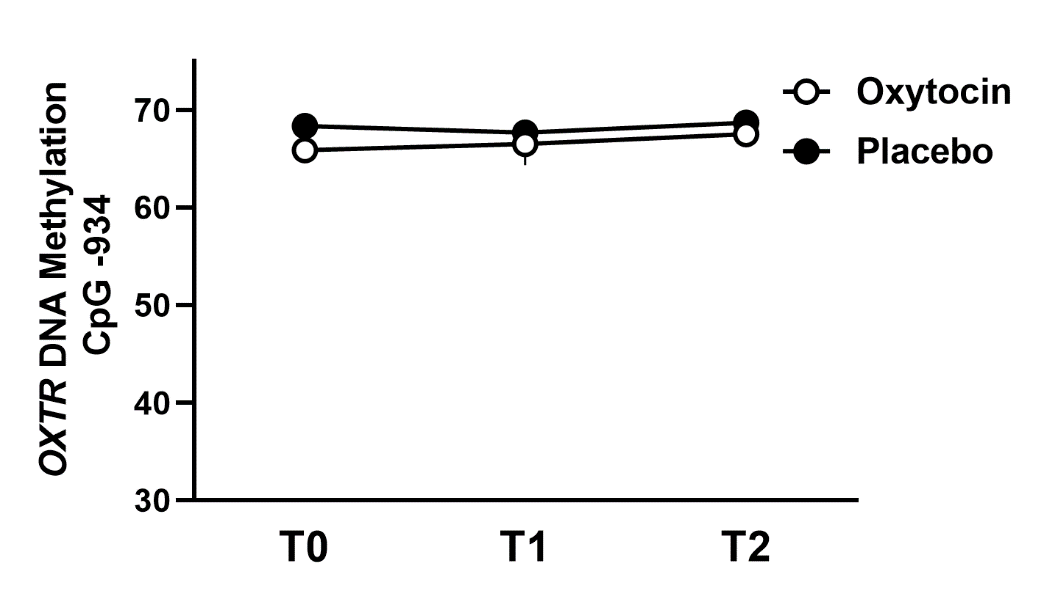

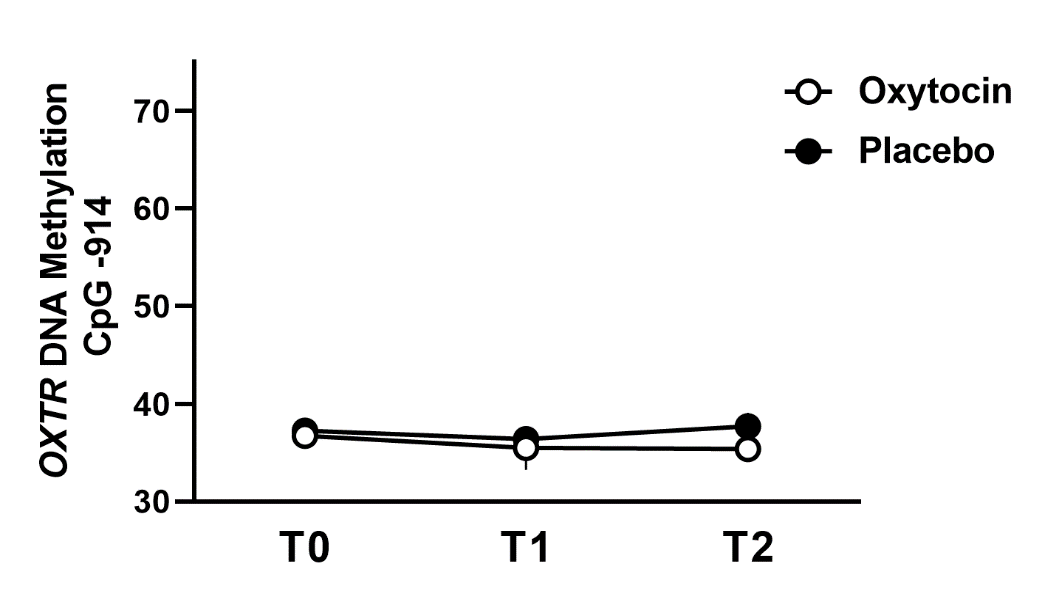

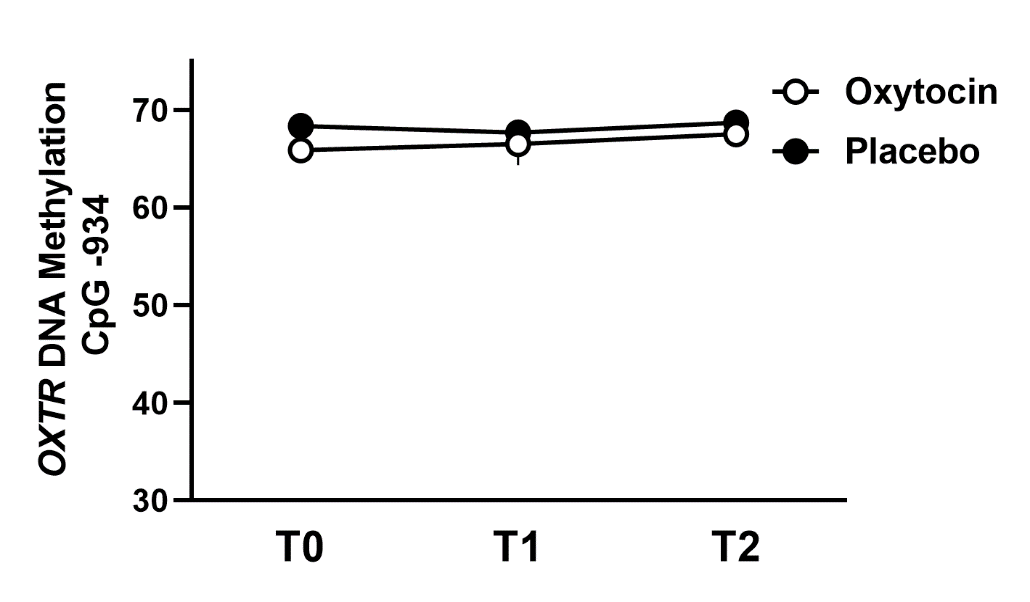

### Supplementary references

Alaerts, K., Daniels, N., Moerkerke, M., Evenepoel, M., Tang, T., Van der Donck, S., Chubar, V., Stephan, C., Steyaert, J., Boets, B., & Prinsen, J. (2023). At the head and heart of oxytocin’s stress-regulatory neural and cardiac effects: a chronic administration RCT in children with autism. *MedRxiv*, 2023.04.04.23288109. https://doi.org/10.1101/2023.04.04.23288109

American Psychiatric Association. (2013). Diagnostic and statistical manual of mental disorders (5th ed.). In *Washington, DC*.

Daniels, N., Moerkerke, M., Steyaert, J., Bamps, A., Debbaut, E., Prinsen, J., Tang, T., Van Der Donck, S., Boets, B., & Alaerts, K. (2023). Effects of multiple-dose intranasal oxytocin administration on social responsiveness in children with autism: a randomized, placebo-controlled trial. *Molecular Autism*, *14*, 16. https://doi.org/10.1186/s13229-023-00546-5

Evenepoel, M., Moerkerke, M., Daniels, N., Chubar, V., Claes, S., Turner, J., Vanaudenaerde, B., Willems, L., Verhaeghe, J., Prinsen, J., Steyaert, J., Boets, B., & Alaerts, K. (2022). Endogenous oxytocin levels in children with autism: Associations with cortisol levels and oxytocin receptor gene methylation. *MedRxiv*, 2022.12.15.22283521. https://doi.org/10.1101/2022.12.15.22283521

Guastella, A. J., Hickie, I. B., McGuinness, M. M., Otis, M., Woods, E. A., Disinger, H. M., Chan, H. K., Chen, T. F., & Banati, R. B. (2013). Recommendations for the standardisation of oxytocin nasal administration and guidelines for its reporting in human research. *Psychoneuroendocrinology*, *38*(5), 612–625. https://doi.org/10.1016/J.PSYNEUEN.2012.11.019

Krol, K. M., Puglia, M. H., Morris, J. P., Connelly, J. J., & Grossmann, T. (2019). Epigenetic modification of the oxytocin receptor gene is associated with emotion processing in the infant brain. *Developmental Cognitive Neuroscience*, *37*, 1–8. https://doi.org/10.1016/j.dcn.2019.100648

Luis Royo, J., Hidalgo, M., & Ruiz, A. (2007). Pyrosequencing protocol using a universal biotinylated primer for mutation detection and SNP genotyping. *Nature Protocols 2007 2:7*, *2*(7), 1734–1739. https://doi.org/10.1038/nprot.2007.244
